# Age- and sex- differences in efficacy of treatments for type 2 diabetes: Network meta-analysis of aggregate and individual level data

**DOI:** 10.1101/2024.06.23.24309242

**Authors:** Peter Hanlon, Elaine Butterly, Lili Wei, Heather Wightman, Saleh Ali M Almazam, Khalid Alsallumi, Jamie Crowther, Ryan McChrystal, Heidi Rennison, Katherine Hughes, Jim Lewsey, Robert Lindsay, Stuart McGurnaghan, John Petrie, Laurie A Tomlinson, Sarah Wild, Amanda Adler, Naveed Sattar, David M Phillippo, Sofia Dias, Nicky J Welton, David A McAllister

## Abstract

**Importance:** Sodium glucose cotransporter 2 inhibitors (SGLT2i), glucagon-like peptide-1 receptor analogues (GLP1ra) and dipeptidyl peptidase-4 inhibitors (DPP4i) improve hyperglycaemia and, in the case of SGLT2i and GLP1ra, reduce the risk of major adverse cardiovascular events (MACE) in type 2 diabetes. It is not clear whether efficacy varies by age or sex.

**Objective:** Assess whether age or sex are associated with differences in efficacy of SGL2i, GLP1ra and DPP4i.

**Data sources:** Medline, Embase and clinical trial registries.

**Study selection:** Two independent reviewers screened for randomised controlled trials of SGLT2i/GLP1ra/DPP4i, compared to placebo/active comparator, in adults with type 2 diabetes.

**Data extraction and synthesis:** We sought individual participant data (IPD) all eligible studies. Where IPD were available, we modelled age- and sex-treatment interactions for each trial. Otherwise, we assessed age- sex distributions along with results from aggregate trial data. IPD and aggregate findings were combined in a Bayesian network meta-analysis.

**Main outcome measures:** HbA1c and MACE.

**Results:** We identified 616 eligible trials (604 reporting HbA1c, 23 reporting MACE) and obtained IPD for 75 trials (6 reporting MACE). Mean age was 59.0 (10.7) years and 64.0 (8.6) in HbA1c and MACE trials, respectively. Proportions of female were 43.1% and 44.0% in HbA1c and MACE trials, respectively. SGLT2i reduced HbA1c by 0.5-1.0% overall compared to placebo. This reduction versus placebo was attenuated in older participants (change in HbA1c 0.25 percentage-points less for 75-year-olds compared to 45-year-olds). SGLT2i showed greater relative efficacy in MACE risk reduction among older than younger people. This finding was sensitive to the exclusion of one of the IPD MACE trials, however, in all sensitivity analyses, SGLT2i were either as efficacious or more efficacious in older participants. There was no consistently significant difference in efficacy by age for GLP1ra or DPP4i for HbA1c or MACE, nor were there consistent significant sex differences for any class.

**Conclusion:** Newer glucose-lowering drugs are efficacious across age and sex groups. SGLT2i are more cardioprotective in older than younger people despite smaller HbA1c reductions. Age alone should not be a barrier to treatments with proven cardiovascular benefit providing they are well tolerated align with patient priorities.

## Introduction

Over the past two decades, newer glucose lowering agents have transformed the management of type 2 diabetes. The efficacy of agents such as SGLT2 inhibitors (SGLT2i) and GLP1 receptor agonists (GLP1ra) in reducing cardiovascular and kidney outcomes is now well established,^1,2^ leading to their widespread use in clinical practice and prominence in clinical guidelines.^3^ Questions remain, however, over how to optimise treatment decisions for individuals, including how characteristics such as age and sex might influence treatment recommendations.^4–6^

Almost half of those with type 2 diabetes are aged over 65 years.^5^ Moreover, age-related functional limitations and conditions such as frailty typically manifest earlier in people with type 2 diabetes.^7^ The risk of complications of diabetes increases with age, potentially increasing the absolute benefits of treatment. Conversely, older adults may also be more susceptible to harms of intensive glycaemic targets.^8,9^ Among women, absolute risk of type 2 diabetes and cardiovascular disease are lower than in men, but diabetes is associated with a greater relative increase in cardiovascular risk in women than men.^10,11^ Women also have different patterns of cardiovascular complications and less intensive management of cardiovascular risk factors from men.^12^ It if therefore important for clinicians, patients and policy makers to understand whether and how responses to treatment differ by age and sex.

Clinical guidelines do not currently recommend different treatments for men and women, nor across different age-groups. They have, however, highlighted the uncertainty that comes from the under-representation of women and older people within trials.^3,13^

Addressing this uncertainty is challenging. Analyses of variation in treatment effects using individual trials (even where there is access to individual-level data) often lacks statistical power.^14^ Although meta-analyses increase power to detect differences in efficacy across participant characteristics, they can produce misleading results due to aggregation bias, that is, making improper inferences about units of analysis (the individual patient) that were not actually analysed.^15^ Individual participant data (IPD) meta-analyses, which pool individual-level data from multiple trials, largely overcome both these deficiencies, but due to legal and practical constraints it is rarely possible to obtain IPD for all relevant trials, limiting gains in power and increasing the risk of selection bias.

We aimed to address these problems via a systematic review and meta-analysis of both aggregate and IPD trial data. Using a meta-analysis technique not susceptible to aggregation bias, we estimated whether the efficacy of newer glucose lowering medications for type 2 diabetes differs according to age and sex.

## Methods

This systematic review and network meta-analysis followed a prespecified protocol (PROSPERO:CRD42020184174).^16^ The protocol covers a wider project for calibration of the network meta-analysis to a community sample. This manuscript presents findings from the assessment of age- and sex-treatment interactions (prior to calibration). Findings are reported according to Preferred Reporting In Systematic Reviews and Meta-analyses (PRISMA) guidelines.^17^

## Eligibility criteria and search strategy

Eligible studies were randomised trials that recruited adults (≥18 years) with type 2 diabetes, and which assessed efficacy of either SGLT2i, GLP1ra, or DPP4 inhibitors (DPP4i) on either glycated haemoglobin (HbA1c) or major adverse cardiovascular events (MACE, defined as death from cardiovascular causes, non-fatal myocardial infarction or non-fatal stroke) compared with either placebo or an active comparator of any other drug class. We excluded within-class comparisons and trials that were not registered.

We searched two electronic databases (Medline and Embase) using both keywords and Medical Subject Headings (full search terms shown in the appendix) as well as the United States and Chinese clinical trial registries. All titles and abstracts were screened, retaining all potentially eligible studies for full text review. All stages of screening were completed by two reviewers working independently, with conflicts resolved by consensus and involving a third reviewer where required.

For all eligible trials, we assessed whether IPD were available for analysis by third party researchers through the Vivli repository, and applied to the independent steering committee for access where this was the case.

### Data extraction

Drug names, doses and regimens were extracted from text strings obtained from clinicaltrials.gov and published documents (papers and clinical study reports). Age and sex at baseline were obtained from published documents. HbA1c results were extracted from clinicaltrials.gov where available or published documents if not. For MACE, results were obtained via manual extraction from published documents (including age- and sex-subgroups). For IPD trials, data were cleaned and harmonised in the Vivli repository.

We assessed risk of bias in each study using the Cochrane Risk of Bias tool.^18^

### Statistical analysis

Detailed description of the statistical analysis is in the supplementary appendix, and summarised briefly here. All data (IPD-summaries and aggregate level) are available at the project github repository https://github.com/Type2DiabetesSystematicReview/nma_agesex_public.

First, we summarised the age- and sex-distribution for each trial using IPD where available, or from published summary statistics otherwise. Then, for each outcome (HbA1c and MACE) we fitted multilevel network meta-regression (ML-NMR) models using the multinma package in R.^19^ These models are described in more detail in our protocol paper and in more technical detail in the original methods paper.^16,19^ This modelling approach was chosen as i) it does not disrupt randomisation ii) it makes less stringent assumptions than standard NMA and iii) it can (without causing aggregation bias) accommodate IPD, aggregate-level trial data and subgroup-level trial data in models estimating treatment-covariate interactions.

For HbA1c, we separately fit network meta-analyses for trials of mono-, dual- and triple-therapy, reflecting different indications for the drugs in question. All MACE trials were in people already established on treatment. We dropped treatment arms evaluating the combined effect of two or more treatments. For the SGLT2i, GLP1ra, DPP4i and metformin, treatment arms were uniquely identified using drug and dose. Insulin, in which dose is titrated, was modelled as a single category. For the remaining drug classes, arms within the same trial with different doses but the same drug were collapsed into a single arm. For all models, placebo was the reference treatment.

In trials for which we had IPD, we fitted trial-level regression models of each outcome by age, sex and treatment, as well as assessing age-treatment and sex-treatment interactions. For HbA1c we fitted linear regression models (additionally including HbA1c at baseline as a covariate). Where participants did not complete the trial we carried forward the last recorded value. For the MACE outcomes we fitted Cox regression models. We checked proportional hazards assumption by plotting scales Schofield residuals. These IPD estimates were then meta-analysed along with aggregate trial-level and (for MACE) subgroup-level data on trial outcomes and on the age- and sex-distributions of each trial (as described in the supplementary appendix). Placebo was used as the reference category in all models. Models were summarised using the posterior mean and 95% credible interval for both the main effect and both age-treatment and sex-treatment interactions.

## Results

### Systematic review results

We identified 685 eligible trials, of which 616 were included in the network meta-analyses (Figure 1). There were a total of 541 aggregate level and 75 IPD trials, including 230,608 and 75,850 participants, respectively. Trial-level details and risk of bias are shown in the online repository.

**Figure 1:**
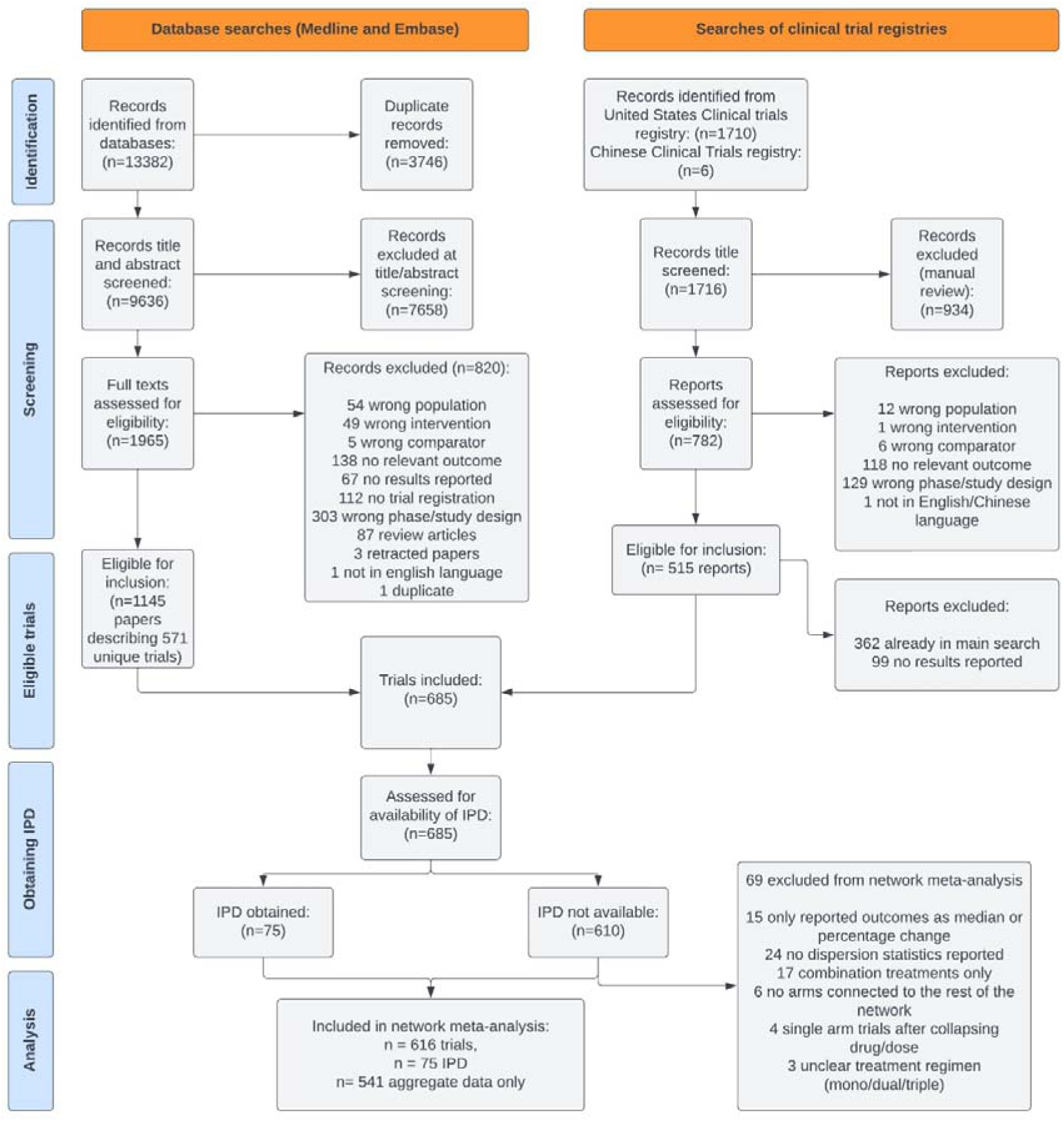
PRISMA diagram of included studies.

Table 1 shows the total number of included trials reporting HbA1c for each class along with aggregate baseline characteristics. Characteristics were similar for trials with IPD and those with aggregate trials. For trials reporting MACE, trial-level details are shown in table 2. For both HbA1c and MACE reporting trials, almost all trial participants were in the 40 to 80 year age range, including trials targeted at older people (supplementary figures S1). For trials reporting HbA1c where IPD was available, age was similarly distributed.

**Table 1:** Trials reporting HbA1c, comparisons and characteristics.

| Classes | Dipeptidyl peptidase 4 inhibitors |  | Glucagon-like peptide-1 analogues |  | Sodium-glucose co-transporter 2 inhibitors |  | Total trials |  |
| --- | --- | --- | --- | --- | --- | --- | --- | --- |
|  | Aggregate | IPD | Aggregate | IPD | Aggregate | IPD | Aggregate | IPD |
| Total | 252 | 33 | 178 | 20 | 150 | 29 | 530 | 74 |
| Placebo | 128 | 22 | 84 | 10 | 102 | 18 | 309 | 47 |
| **Specific drugs of the following classes** |  |  |  |  |  |  |  |  |
| Dipeptidyl peptidase 4 (DPP-4) inhibitors | - | - | 20 | 2 | 19 | 6 | 284 | 41 |
| Glucagon-like peptide-1 (GLP-1) analogues | 27 | 2 | - | - | 9 | 0 | 246 | 32 |
| Sodium-glucose co-transporter 2 (SGLT2) inhibitors | 20 | 9 | 9 | 0 | - | - | 183 | 51 |
| Sulfonylureas | 27 | 4 | 8 | 1 | 12 | 3 | 46 | 7 |
| Biguanides (metformin only) | 24 | 9 | 4 | 1 | 3 | 3 | 30 | 13 |
| Thiazolidinediones | 15 | 0 | 5 | 1 | 4 | 0 | 22 | 1 |
| Alpha glucosidase inhibitors | 12 | 1 | 2 | 0 | 1 | 0 | 14 | 1 |
| 'Other blood glucose lowering drugs, excl. insulins', eg repaglinide | 2 | 0 | 3 | 0 | 0 | 0 | 4 | 0 |
| **Any drug of the following class** |  |  |  |  |  |  |  |  |
| Insulins and analogues (eg "any insulin") | 5 | 0 | 42 | 6 | 1 | 0 | 46 | 6 |
| Blood glucose lowering drugs, excl. insulins (eg "any oral antidiabetic drug") | 1 | 0 | 3 | 0 | 0 | 0 | 4 | 0 |
| 2 arms | 219 | 18 | 124 | 10 | 115 | 10 | 427 | 34 |
| 3 arms | 25 | 9 | 43 | 7 | 28 | 15 | 82 | 30 |
| 4 or 5 arms | 8 | 6 | 11 | 3 | 5 | 4 | 21 | 10 |
| Participants | 111580 | 27563 | 88302 | 21280 | 44794 | 33277 | 229115 | 75850 |
| Male n (%) | 64333<br>(57.7%) | 15414<br>(55.9%) | 49179<br>(55.7%) | 12707<br>(59.7%) | 25272<br>(56.6%) | 19727<br>(59.3%) | 130154<br>(56.9%) | 44557<br>(58.7%) |
| Age, years (sd) [5<sup>th</sup> to 95<sup>th</sup><br>centile] | 58.7 (10.8)<br>[40.3-75.8] | 57.2 (11.1)<br>[36.8-74.9] | 57.7 (10.3)<br>[40.4-74.2] | 60.0 (10.8)<br>[41.1-76.4] | 61.2 (10.7)<br>[42.7-78.0] | 56.8 (11.3)<br>[35.2-74.6] | 59.0 (10.7)<br>[40.9-75.9] | 57.9 (11.2)<br>[37.1-75.3] |
| The number of trials in each class do not sum to the total because some trials include more than one class. |  |  |  |  |  |  |  |  |

**Table 2:**
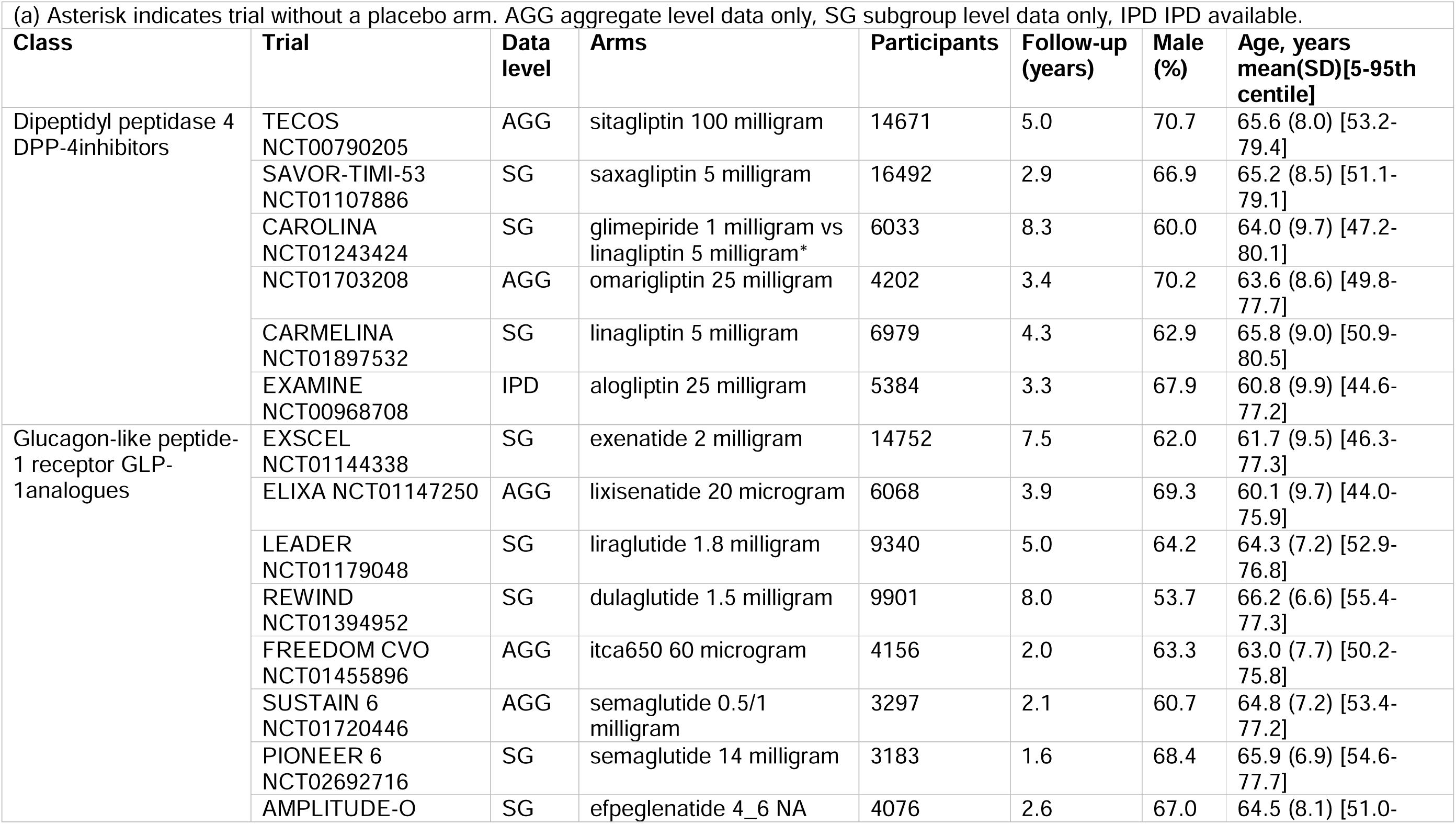
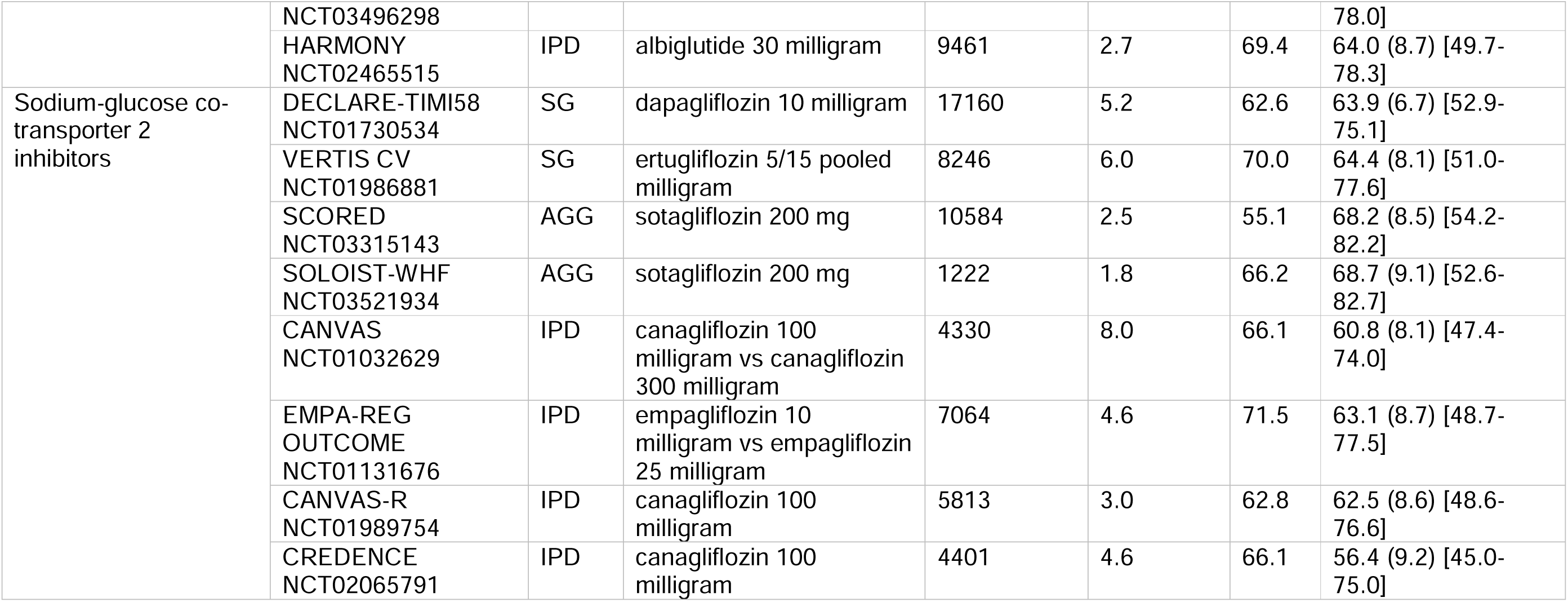
MACE Trials, characteristics.

### Main treatment effects

The main treatment effects comparing each treatment versus placebo are shown in supplementary figure S2 for HbA1c for a standard network meta-analysis without covariates. Most treatments reduced HbA1cwith a range of absolute reductions of −0.5% to −1.5%.

Supplementary figure S3 shows the same analysis for MACE, showing the expected result of reduced hazard of MACE for SGLT2i compared to placebo, with null findings for DPP4i and for some agents within the GLP1ra class (consistent with trial-level findings for these agents which did not show superiority to placebo).

### Age-treatment and sex-treatment interactions

Figure 2 shows the age-treatment and sex-treatment interactions, assessing differences in the efficacy of treatment by age and sex, for HbA1c (panel a) and MACE (panel b). For HbA1c, interactions are on the absolute scale showing change in HbA1c in units of percent. Across all three networks, SGLT2-inhibitors were less efficacious with increasing age; a 30-year increment in age was associated with an attenuation in effect on Hba1c (%) of around 0.25 percentage points on the absolute scale (equivalent to about a quarter to half of the overall treatment effect). There was also some evidence of a sex-treatment interaction for HbA1c across networks for SGLT-2 inhibitors, although this was small in magnitude (around 0.05 percentage points) and the upper range of the credible intervals included the null. No consistent differences across networks were apparent for any other age-treatment or sex-treatment interactions.

**Figure 2:**
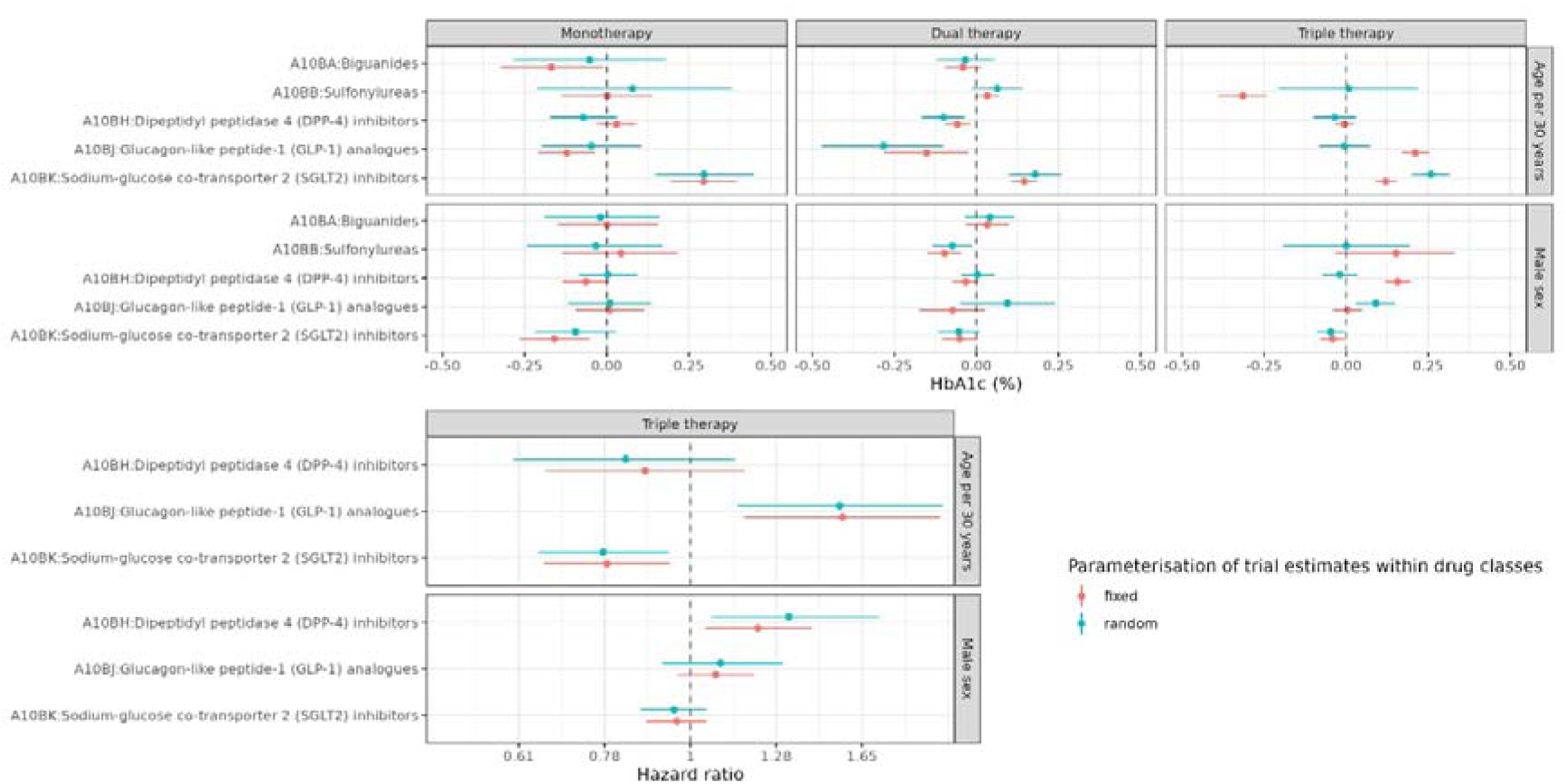
Covariate-treatment interactions for HbA1c and MACE: This figure shows the covariate-treatment interaction estimates for age and sex represented as dots, both for a) HbA1c (top panels) and b) MACE (bottom panel). Horizontal lines show the 95% credible interval. Age was modelled as a continuous variable and divided by 30 (so that the coefficient reflects the difference in efficacy over a 30-year age difference). Estimates below the line of no effect (dashed vertical line) indicate that the treatment is more efficacious in older age/in male sex. Estimates above this line indicate the inverse.

For MACE, there was evidence of greater relative efficacy in older people for SGLT-2i. For GLP-1ra, there was some evidence of lower efficacy in older people. There was also some evidence of an age-treatment interaction for DPP-4 inhibitors, with higher efficacy in older people (however the main effect in this class was null). When modelling sex-treatment interactions in MACE trials, DPP-4i appeared to be less efficacious in men, although this association was less evident in trials which included sex-subgroup data and absent on excluding the sole DPP-4i trial for which we had access to IPD. For GLP-1ra and SGLT-2i there was no evidence of any sex-treatment interactions.

In sensitivity analyses including/excluding age- and sex-subgroup data in the model, and dropping each trial in turn, the greater efficacy in older people for SGLT2i persisted whether or not age-subgroup data were included in the modelling, but was not found after excluding one of the four SGLT2i IPD trials (NCT01131676, EMPA-REG OUTCOME, see supplementary figure S4). Less equivocally, for older people, despite the finding of attenuated effects of SGLT-2i on HbA1c the reduction in the MACE risk was preserved or greater in all of the sensitivity analyses.

Particular caution is needed in interpreting the GLP-1ra and DPP-4i findings for MACE, as results differed depending on the inclusion/exclusion of the single trials of each agent for which we had IPD (HARMONY NCT02465515 and EXAMINE NCT00968708 for GLP-1ra and DPP4i respectively) and/or the inclusion/exclusion of subgroup data (Supplementary figure S4).

### Age and sex-specific effects for MACE trials

Figure 3 shows the impact of the age-treatment and sex-treatment interactions on the overall age- and sex-specific relative efficacy versus placebo for each class.

**Figure 3:**
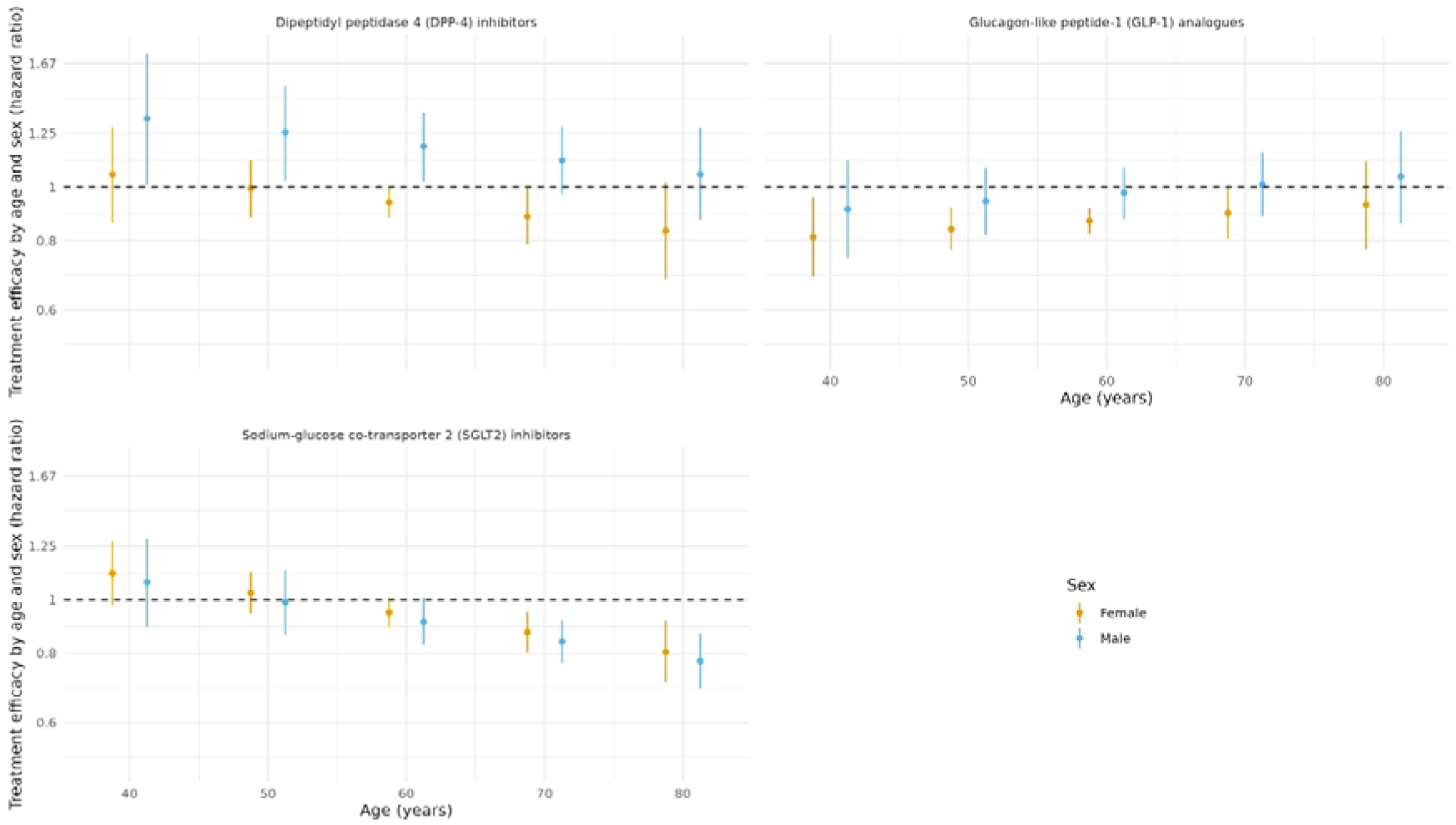
Relative effects for MACE: This figure is based on a model including all available trials, including sex-subgroup data as well as aggregate data and IPD. It shows age- and sex-specific estimates of the effect of each treatment compared to placebo on the hazard of MACE.

Consistent with the findings reported above for DPP-4 inhibitors (that they were not efficacious overall compared to placebo, but with some evidence of differential effect by age and sex), DPP4i were associated with increased risk of MACE in men and younger women but decreased risk in older women. GLP-1ra (which again were inefficacious overall but with differential efficacy by sex) were not associated with significant reduction in MACE in men and in older people, but were associated with decreased risk of MACE in younger women. SGLT2i were associated with lower MACE in older people regardless of sex with equivocal findings for people younger than 60 years. For all three classes, however, some caution is needed in interpreting findings given the sensitivity of the interaction estimates to the inclusion of particular trials. Less equivocally, SGLT-2i were associated with reduced MACE in older men and older women.

## Discussion

This network meta-analysis of 616 trials, including IPD from 75 trials, assessed whether the efficacy of three newer drug classes (SGLT2i, GLP1ra and DPP4i) varied by age or sex in people with type 2 diabetes. For HbA1c, SGLT2i showed modestly reduced efficacy with increasing age, with attenuation of the treatment effect compared to placebo by approximately one quarter at 75 compared with 45 years of age. In contrast, the reduction in MACE with SGLT2i was greater in older compared to younger people. This finding was sensitive to the exclusion of IPD from the EMPAREG-OUTCOME study, however even after excluding this the relative cardiovascular efficacy of SGLT2i preserved for older people. For GLP1ra there was some evidence that cardiovascular efficacy was greater among younger women, but with much greater uncertainty than that seen for SGLT2i.

Previous studies assessing heterogeneity in efficacy, that is, interaction, of type 2 diabetes treatment by age or sex have generally used aggregate or subgroup data from randomised controlled trials, or relied on observational (i.e., non-randomised) data. A meta-analysis of differences between men and women in the efficacy of SGLT2i and GLP1ra found no statistically significant difference in efficacy for cardiovascular outcomes but speculated on possible reduced cardiovascular efficacy among women due to the greater statistical uncertainty in the estimates for this group.^4^ Our analysis, including a larger and more comprehensive group of studies and incorporating IPD, provides greater precision and more clearly demonstrated that sex is not associated with any difference in the efficacy of these classes of medication.

A recent network meta-analysis assessed the efficacy of type 2 diabetes treatment across a range of clinical outcomes, including outcomes such as heart failure, end-stage kidney disease, and medication related-harms that we did not include in this analysis.^2^ This recent network meta-analysis showed that, in addition to MACE, SGLT2i and GLP1ra reduce the risk of admission to hospital with heart failure and of end-stage kidney disease, with SGLT2i showing superior efficacy for the latter. Harms with treatment were generally class-specific and included genital infections with SGLT2i and gastrointestinal complications with GLP1ra. This previous analysis, however, did not assess heterogeneity by age and sex, and did not include analysis of IPD.

One likely explanation for the reduction in glycaemic efficacy of SGLT2i with older age is age-related decline in kidney function. For example, the recent TriMaster study (a double-blind three-way crossover study comparing DPP4i with SGLT2i) demonstrated that participants with estimated glomerular filtration rates 60-90 ml/min/1.73m^2^, compared to those >90 ml/min/1.73m^2^, had lower HbA1c on DPP4 inhibitors than SGLT2 inhibitors.^20^ In this context, it is notable that the reductions in MACE with SGLT2i were greater in older people, despite lower glycaemic efficacy. This highlights the limitation of surrogate outcomes such as HbA1c in determining the risks of harder endpoints such as MACE for which hyperglycaemia is a less important risk factor than hypertension or dyslipidaemia.^21^ The findings also suggest SGLT2i are working to prevent hard outcomes by pathways largely unrelated to conventional risk factors or dysglycemia. Current clinical guidelines recommend less stringent glycaemic targets in older people living with multiple long-term conditions or frailty due to greater risks of adverse events.^3,22^ While caution around pursuing surrogate markers is well founded, this may also lead to clinicians not initiating therapies with other (e.g. cardiovascular) benefits who may have potential to benefit. Our findings highlight the need to also consider cardioprotective effects agents such as SGLT2i (in addition to safety, tolerability and patient’s priorities) when agreeing optimal treatment strategies for older people.

The primary strength of this analysis is in the use of individual participant data to estimate age- and sex-treatment interactions. This improves statistical power and allows integration of individual participant data and aggregate data within network meta-analysis to preserve randomisation and avoid aggregation bias. We also followed rigorous systematic review methodology to identify eligible studies, and have made all model outputs and analysis code publicly available to facilitate replication of our findings. However, despite the inclusion of a large volume of IPD, this was not available for all trials. Furthermore, the trials for which we did have IPD were not a random sample of the included trials as their availability depended on the data sharing arrangements put in place by the sponsor. We dropped trial arms with multiple drug classes as the software does not allow for explicit modelling of components within arms, and our focus was on class-level interactions. While we assessed glycaemic and cardiovascular efficacy, which are clinically relevant outcomes, our analysis did not include other clinical endpoints (such as kidney events) or analysis of adverse events and safety, which would be relevant outcomes for future analyses.

While our findings demonstrate similar or better cardiovascular efficacy among older people within these trials, trials rarely enrol people over 80 years of age. Furthermore, age-associated states such as frailty, which increase the risk of both cardiovascular events and also of complications,^8,23^ are not quantified in these trials. This analysis does not, therefore, assess whether efficacy is similar in people of much higher ages (i.e. over 80 years) or living with frailty. This is a group in which the balance of risks and benefits is most uncertain, and as such several complementary strands of evidence are needed. Retrospective assessment of frailty within trial IPD (e.g. using the cumulative deficit frailty index) can allow assessment of frailty within trial participants,^24^ but those with most severe frailty are likely to be excluded from the trial altogether. Routine healthcare data may be used to model natural history and rates of complications across a range of ages and health states, allowing modelling of overall benefit. Observational analyses using causal inference methods may also be utilised to assess safety, including in people who may be under-represented in trials. However, biases such as confounding by indication remain a significant challenge in such analyses. Finally, where uncertainty remains, there is a need for trials that recruit and retain older people and those living with frailty, and which explicitly measure and report functional status.

In conclusion, the efficacy of newer glucose lowering agents is broadly consistent between men and women. The glycaemic efficacy of SGLT2i is modestly reduced with increasing age, but cardiovascular event reduction is preserved. Indeed, incorporating analyses of IPD within our meta-analysis demonstrated that the reduction in MACE seen in with SGLT2i versus placebo was greater among older people. Antidiabetic agents are beneficial on a number of end points, and we demonstrate that those benefits may vary substantially and differently with age. Treatment decisions based on glycaemia alone could potentially lead to removal of agents with cardiovascular benefit. Guidelines should emphasise approaches where antidiabetic drugs such as SGLT2i are used to reduce cardiovascular risk as the aim – as agreed with patients – rather than overemphasising glycaemic targets.

## Acknowledgements

Data access and provision: This manuscript is based on research using data from data contributors Lilly, Boehringer Ingelheim, Sanofi, Takeda, GlaxoSmithKline, AstraZeneca and Johnson & Johnson that has been made available through Vivli, Inc. Vivli has not contributed to or approved, and is not in any way responsible for, the contents of this publication.

## Author contributions

PH, EB and DM conceived the study. EB, LW and PH performed the literature search and screened articles for inclusion. EB, PH, LW, JC, HR, SA and KA extracted aggregate data from the included studies. DM, HW, JC, RM and PH accessed and processed the individual-level data. DM wrote the statistical analysis plan with DP, SD and NW providing statistical input. DM performed the analysis with input from DP, SD, NW and PH on analysis outputs. PH wrote the first draft. EB, LW, HW, SA, KA, JC, RM, HR, KH, JL, RL, SM, JP, LT, SW, AA, NS, DP, SD, NW and DM reviewed this and subsequent drafts providing critical input. All authors approved the final version for submission.

## Data availability

Individual-level participant data was obtained through the Vivli project, subject to a data sharing agreement. Data are available on application to the data holder via Vivli’s application process. All aggregate data, as well as summary data from all analyses of individual participant data, are available at https://github.com/Type2DiabetesSystematicReview/nma_agesex_public, along with analysis code for all the analyses presented in the manuscript and supplementary appendices.

## References

1. Palmer SC, Tendal B, Mustafa RA, et al. Sodium-glucose cotransporter protein-2 (SGLT-2) inhibitors and glucagon-like peptide-1 (GLP-1) receptor agonists for type 2 diabetes: systematic review and network meta-analysis of randomised controlled trials. BMJ 2021; 372: m4573.

2. Shi Q, Nong K, Vandvik PO, et al. Benefits and harms of drug treatment for type 2 diabetes: systematic review and network meta-analysis of randomised controlled trials. BMJ 2023; 381: e074068.

3. Davies MJ, Aroda VR, Collins BS, et al. Management of hyperglycemia in type 2 diabetes, 2022. A consensus report by the American Diabetes Association (ADA) and the European Association for the Study of Diabetes (EASD). Diabetes care 2022; 45(11): 2753–86.

4. Singh AK, Singh R. Gender difference in cardiovascular outcomes with SGLT-2 inhibitors and GLP-1 receptor agonist in type 2 diabetes: a systematic review and meta-analysis of cardio-vascular outcome trials. Diabetes & Metabolic Syndrome: Clinical Research & Reviews 2020; 14(3): 181–7.

5. Bellary S, Kyrou I, Brown JE, Bailey CJ. Type 2 diabetes mellitus in older adults: clinical considerations and management. Nature Reviews Endocrinology 2021; 17(9): 534–48.

6. Bellary S, Barnett AH. SGLT2 inhibitors in older adults: overcoming the age barrier. The Lancet Healthy Longevity 2023; 4(4): e127–e8.

7. Hanlon P, Fauré I, Corcoran N, et al. Frailty measurement, prevalence, incidence, and clinical implications in people with diabetes: a systematic review and study-level meta-analysis. The Lancet Healthy Longevity 2020.

8. Nguyen TN, Harris K, Woodward M, et al. The Impact of Frailty on the Effectiveness and Safety of Intensive Glucose Control and Blood Pressure–Lowering Therapy for People With Type 2 Diabetes: Results From the ADVANCE Trial. Diabetes Care 2021; 44(7): 1622–9.

9. Miller ME, Williamson JD, Gerstein HC, et al. Effects of randomization to intensive glucose control on adverse events, cardiovascular disease, and mortality in older versus younger adults in the ACCORD trial. Diabetes Care 2014; 37(3): 634–43.

10. Peters SA, Huxley RR, Woodward M. Diabetes as a risk factor for stroke in women compared with men: a systematic review and meta-analysis of 64 cohorts, including 775 385 individuals and 12 539 strokes. The Lancet 2014; 383(9933): 1973–80.

11. Peters SA, Huxley RR, Woodward M. Diabetes as risk factor for incident coronary heart disease in women compared with men: a systematic review and meta-analysis of 64 cohorts including 858,507 individuals and 28,203 coronary events. Diabetologia 2014; 57: 1542–51.

12. Peters SA, Woodward M. Sex differences in the burden and complications of diabetes. Current diabetes reports 2018; 18: 1–8.

13. Clemens KK, Woodward M, Neal B, Zinman B. Sex disparities in cardiovascular outcome trials of populations with diabetes: a systematic review and meta-analysis. Diabetes Care 2020; 43(5): 1157–63.

14. Brookes ST, Whitely E, Egger M, Smith GD, Mulheran PA, Peters TJ. Subgroup analyses in randomized trials: risks of subgroup-specific analyses;: power and sample size for the interaction test. Journal of clinical epidemiology 2004; 57(3): 229–36.

15. Geissbühler M, Hincapié CA, Aghlmandi S, Zwahlen M, Jüni P, da Costa BR. Most published meta-regression analyses based on aggregate data suffer from methodological pitfalls: a meta-epidemiological study. BMC Medical research methodology 2021; 21(1): 123.

16. Butterly E, Wei L, Adler AI, et al. Calibrating a network meta-analysis of diabetes trials of sodium glucose cotransporter 2 inhibitors, glucagon-like peptide-1 receptor analogues and dipeptidyl peptidase-4 inhibitors to a representative routine population: a systematic review protocol. BMJ open 2022; 12(10): e066491.

17. Page MJ, McKenzie JE, Bossuyt PM, et al. Updating guidance for reporting systematic reviews: development of the PRISMA 2020 statement. Journal of Clinical Epidemiology 2021; 134: 103–12.

18. Higgins J, Altman D, Sterne J. Assessing risk of bias in included studies. In: Higgins J, Churchill R, Chandler J, Cumpston M, eds. Cochrane Handbook for Systematic Reviews of Interventions version 520 (updated June 2017): Cochrane.

19. Phillippo DM. multinma: An R package for Bayesian network meta-analysis of individual and aggregate data. Evidence Synthesis and Meta-Analysis in R Conference 2021; 2021; 2021.

20. Shields BM, Dennis JM, Angwin CD, et al. Patient stratification for determining optimal second-line and third-line therapy for type 2 diabetes: the TriMaster study. Nat Med 2023; 29(2): 376–83.

21. Yudkin JS, Lipska KJ, Montori VM. The idolatry of the surrogate. Bmj 2011; 343.

22. Strain WD, Down S, Brown P, Puttanna A, Sinclair A. Diabetes and frailty: an expert consensus statement on the management of older adults with type 2 diabetes. Diabetes Therapy 2021; 12: 1227–47.

23. Hanlon P, Jani BD, Butterly E, et al. An analysis of frailty and multimorbidity in 20,566 UK Biobank participants with type 2 diabetes. Communications Medicine 2021; 1(1): 1–9.

24. Hanlon P, Butterly E, Lewsey J, Siebert S, Mair FS, McAllister DA. Identifying frailty in trials: an analysis of individual participant data from trials of novel pharmacological interventions. BMC Med 2020; 18(1).

