## Supplementary appendix for "Age- and sex- differences in efficacy of treatments for type 2 diabetes: Network meta-analysis of aggregate and individual level data"

This supplementary appendix contains a more detailed description of the study methods along with further results as detailed and referenced within the main text.

All extracted data from aggregate studies (trial characteristics, baseline characteristics and results), all model outputs from analyses of individual-level trial data, risk of bias assessments, and analysis code for all results presented within the main paper and the supplementary appendix can be found at <https://github.com/Type2DiabetesSystematicReview/nma_agesex_public>.

### Supplementary methods

#### Search strategy

Full search terms used in each database are shown in <https://bmjopen.bmj.com/content/bmjopen/12/10/e066491.full.pdf?with-ds=yes>

#### Data extraction

WHO ATC drug names, drug doses and regimens were extracted from text strings obtained from clinicaltrials.gov and published documents (papers and clinical study reports). HbA1c results were extracted from clinicaltrials.gov where available or clinical documents if not. Outcomes were captured at arm, contrast and/or pre-and post intervention level. Age and sex at baseline were obtained from published documents, first by reading the tables into software for processing tabular data (https://www.tabletidier.org) then into R. All results were checked manually. For MACE, results were obtained via manual extraction from published documents. For IPD trials, data were cleaned and harmonised in the Vivli repository.

#### Statistical analysis

All analyses requiring access to IPD were conducted within the Vivli safe haven/trusted research environment (hereafter, Vivli). All other analyses were conducted on machines within the University of Glasgow (hereafter, local). All analyses involving IPD were conducted in two stages. First, trial-level analyses of IPD were produced within Vivli and summaries were exported. Secondly, these exported summaries were combined locally with aggregate level data. This two-step approach was chosen to meet the terms of the data sharing agreement, minimise the use of intensive computing resources within the Vivli environment, and to maximise data reuse and ensure reproducibility.

##### Baseline characteristics

In five of the IPD MACE trials, age was plotted within Vivli and found to be well-approximated with a normal distribution. Therefore, these were summarised using the mean and standard deviation. In the remaining MACE trial, age had been binned into three categories during the anonymisation process before it was provided to us, so the number of participants in each age category was summarised using simple counts. Sex was summarised as the number of participants in each trial and arm who were male and female.

As age in some of the IPD HbA1c trials did not follow a normal distribution, we summarised the distributions using empirical cumulative distribution functions (ECDFs). We estimated the ECDFs within each arm and sex and summarised these using the Ramer–Douglas–Peucker algorithm (https://cran.r-project.org/package=RDP), which reduces the number of points required to represent a curve or polygon. As a check on the summaries, they were plotted against the full ECDFs before being exported from Vivli. In stage two, we recreated full ECDFs by linear interpolation, and sampled from these to obtain age distributions.

For aggregate-level trials (both HbA1c and MACE) we estimated the age distribution from published summary statistics. For trials with age cut-offs below 20 years and above 100 years we assumed a normal distribution, otherwise we assumed a truncated normal distribution. We estimated the central tendency parameter ($\mu$) and dispersion parameter ($\sigma$) of the latter by numerical optimisation, based on the reported mean, standard deviation and upper and lower age limits. As, based on the IPD trials, age was only weakly associated with sex, we assumed the age distribution was the same for men and women. To obtain plots and summary statistics for the age distributions across trials we sampled from truncated normal/normal distributions setting the number of samples equal to the number of participants in each arm/sex stratum. As a check on this approach we repeated this method for the IPD trials comparing the results to the ECDF-derived distributions described above (see Figure S 1 (a)).

##### Multi-level network meta-regression models (ML-NNR)

Each model was first fitted assuming identical (fixed) effects across trials for the same treatment. For all HbA1c analyses and selected MACE analyses, this assumption was then relaxed to allow exchangeable (random) effects. Main effects for the covariates and covariate-treatment interactions were assumed to be common across trials within the same drug class.

For IPD, data can be included either as the raw data or, equivalently, as outputs from trial-level models fitted to the data (ie the coefficients and variance-covariance matrices). We implemented the latter approach in the multinma package. Within Vivli, for each of the IPD trials, we fitted trial-level regression models of each outcome on age, sex and treatment as well as age-treatment and sex-treatment interactions. For HbA1c we fitted linear regression models (additionally including HbA1c at baseline as a covariate). For the MACE outcomes we fitted Cox regression models. In both cases we exported the model outputs from the Vivli environment.

For HbA1c aggregate-level trials, the outcomes were modelled using arm-level data (the change in HbA1c in each arm and the accompanying standard error), contrast-level data (the difference between arms and standard error) or, rarely, as post-treatment estimates. For MACE aggregate-level trials the outcome was modelled as contrast level estimates (log-hazard ratios and standard errors for log-hazard ratios). For HbA1c, since the outcome is continuous, only the means for the covariates are required (ie the mean age, mean baseline HbA1c and the proportion of men in each trial/arm). As MACE is not modelled on a linear scale the full joint distribution of the covariates (age and sex) was used - the age distributions were represented using truncated normal distributions (obtained as described above) with sex represented as a Bernoulli distribution (with the parameter equal to the percentage of men in each trial arm). The correlation between these variables was assumed to be the same as that observed in trials for which we had IPD.

For MACE trials we also modelled, separately, subgroup data for age and sex alongside the IPD and aggregate level data. For the sex subgroup data, we assumed the same age distribution across sex strata. For the age subgroup data, we assumed the same proportion were male across age strata. As before, age was represented using truncated normal distributions, with the same $\mu$ and $\sigma$ as the earlier models without subgroup data, but with cut-points equal to the stratum-defining age limits (eg µ= 60 $\sigma=10$, lower = 20 and upper = 69 for a trial with an age subgroup cut-point of 70). Finally, in order to estimate age-sex specific efficacy for MACE at drug-class level, we labelled all treatment arms according to the drug class (ignoring specific drug/dose) and re-ran the model with IPD, aggregate level and sex-subgroup data.

For all models we ran four chains for 2,000 iterations each, and checked for divergent transitions, model convergence (visually using caterpillar plots and using the Gelman-Ruben statistic) and autocorrelation using the R shinystan package. These models were run on a high-performance computing environment within the University of Glasgow. See appendix for additional details of modelling and github for the data and multinma model code (https://github.com/Type2DiabetesSystematicReview/nma_agesex_public). The first 1,000 iterations from each chain were discarded, leaving 4,000 samples. Models were summarised using the mean and 95% credible interval, obtained as the 2.5th and 97.5th quantiles of the 4,000 samples.

### Supplementary figures and tables

#### Age distributions

Figure S 1 shows the age distribution results in violin plots (similar to density plots, but turned through 90 degrees so more can be visualised). Figure S 1 (a) also shows the 2.5th and 97.5th quantiles as horizontal lines. Figure S 1 (a) shows all MACE trials, all IPD trials and a random sample of aggregate-level HbA1c trials. Except for IPD HbA1c trials, or trials where older people were explicitly excluded, the tails of the violin plots may overestimate the presence of people over the age of 80 (as they assume no truncation). Data was not available on differences in age by sex for aggregate level trials. For HbA1c trials where IPD was available, age was similarly distributed by sex (Figure S 1 (b)). For MACE IPD trials, the difference in age between men and women was generally around two years Table S 1. The extent to which the truncated normal distributions estimated from summary statistics for these analyses match empirical cumulative distribution functions (ECDFs) is shown in Figure S 1 (c).

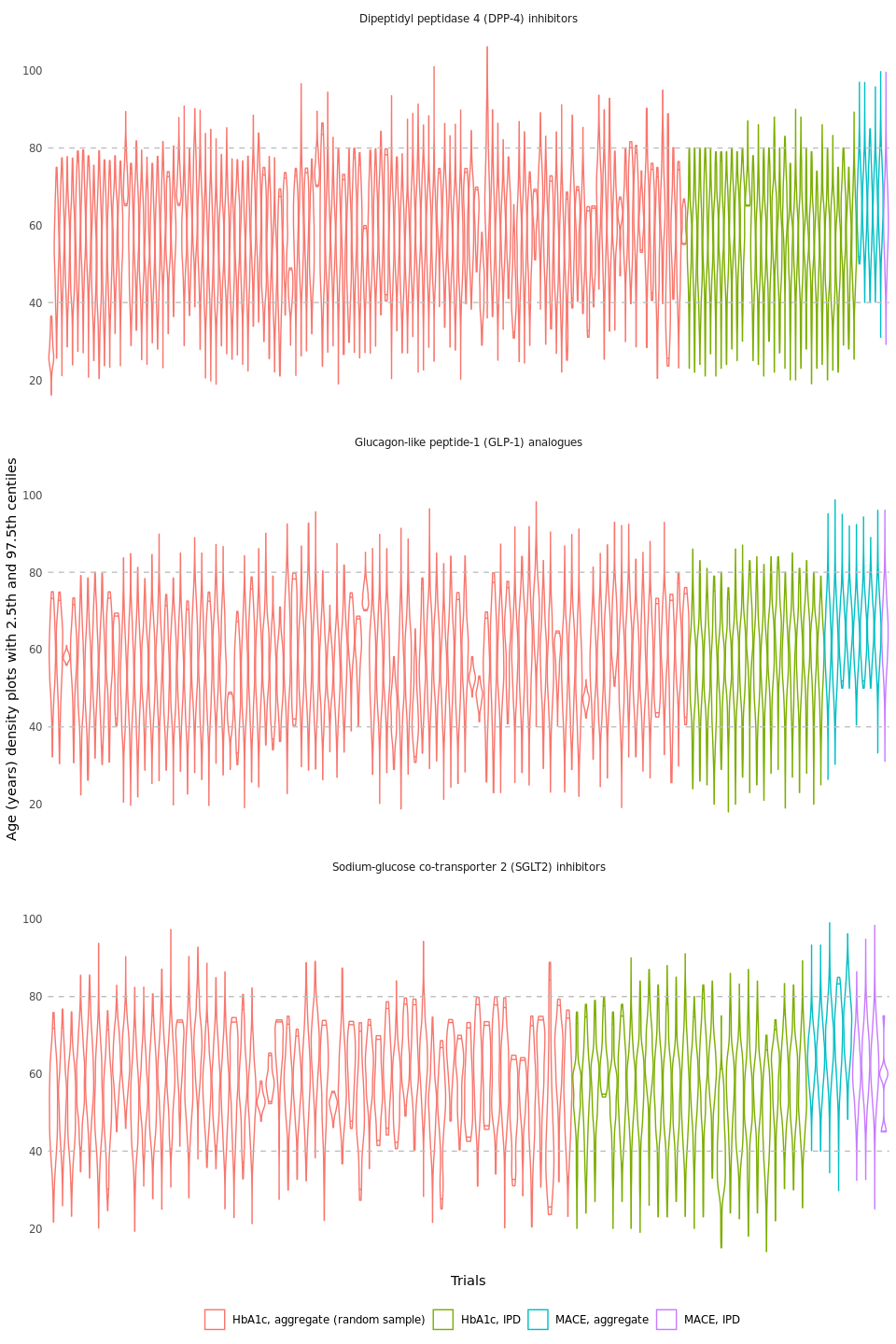

(a)

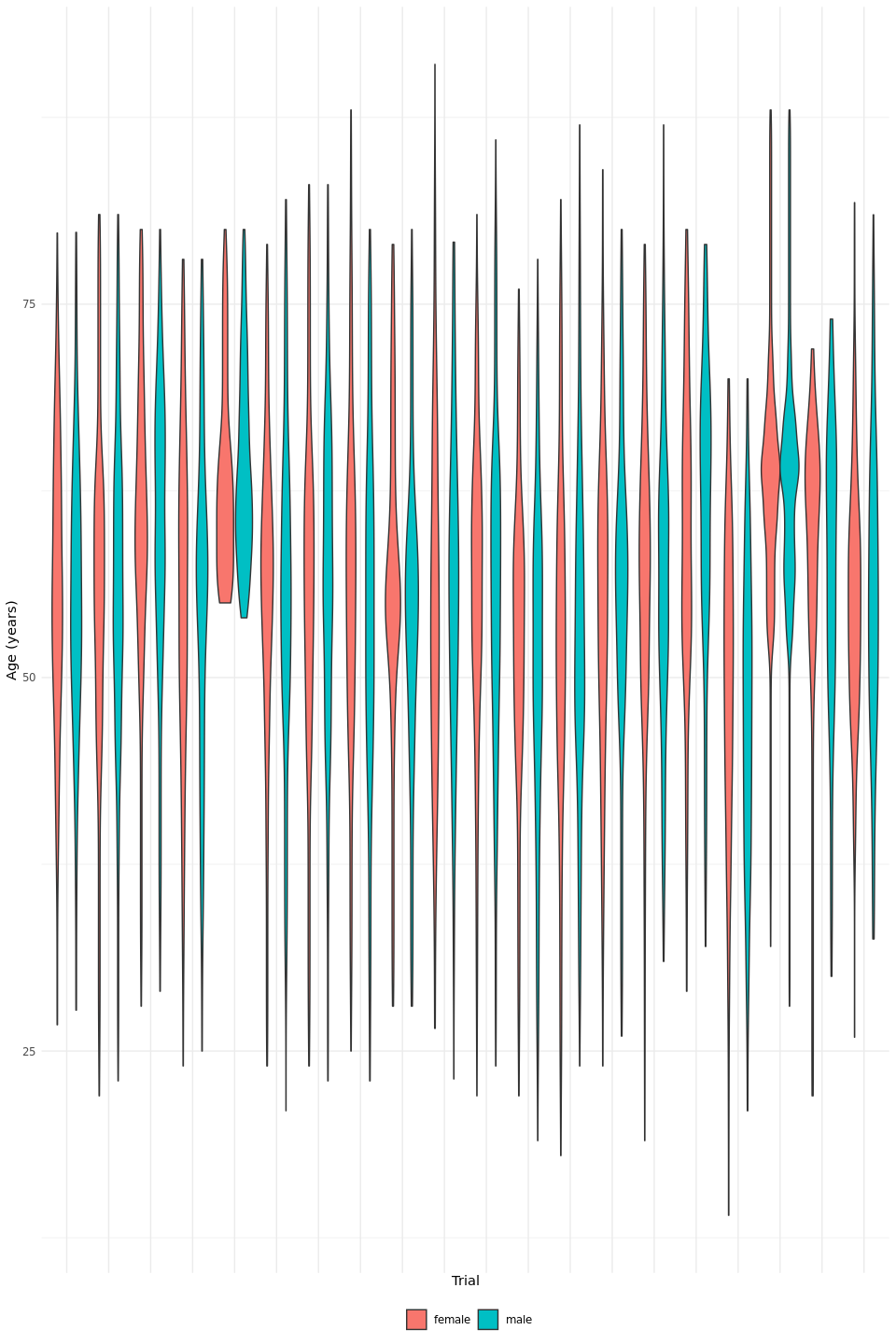

(b)

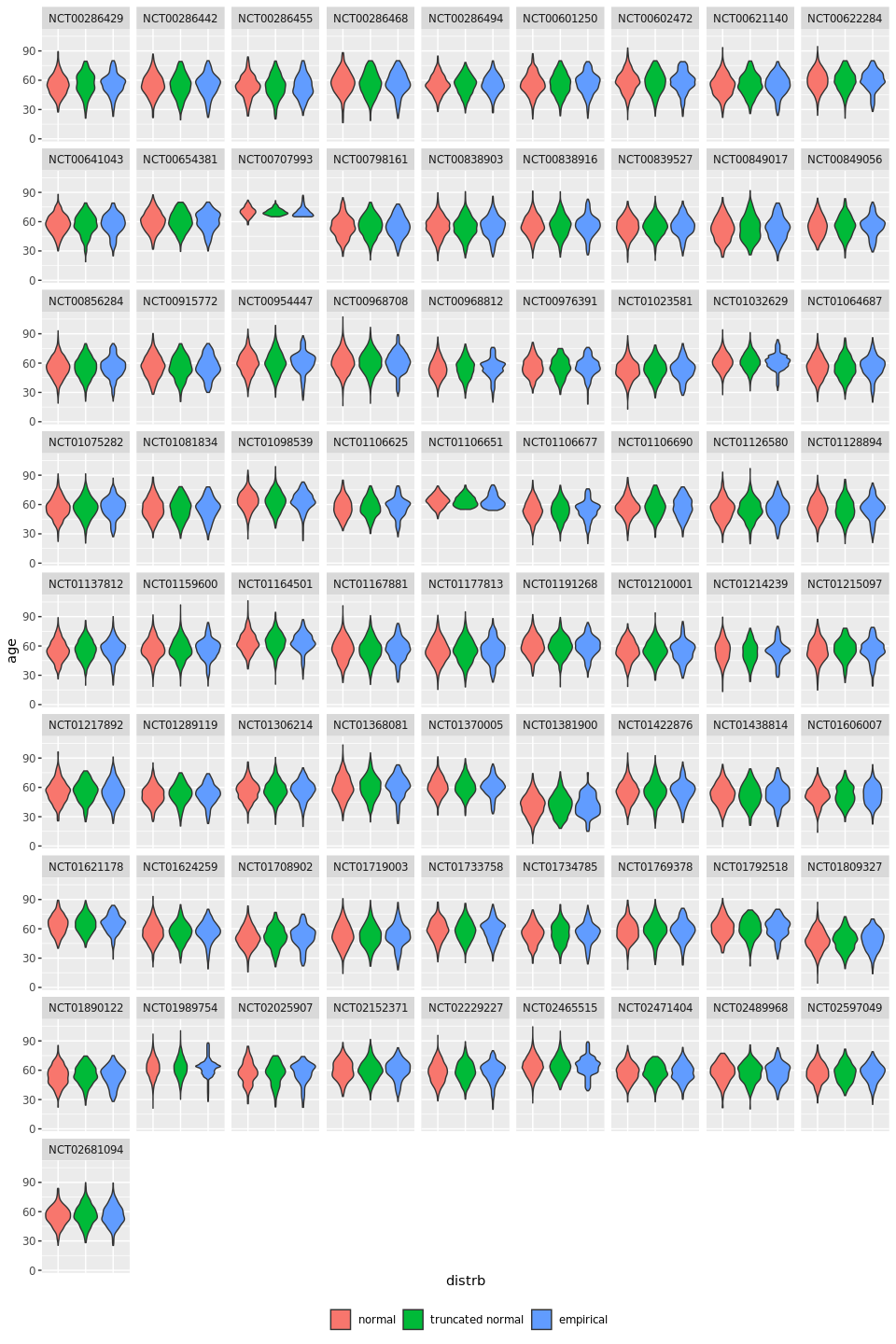

(c)

Figure S 1: Trial age distributions

Table S 1: Mean age in years by sex and arm for MACE trials

| nct_id | arm | F | M | Difference |
| --- | --- | --- | --- | --- |
| NCT00968708 | alogliptin | 65.7 | 63.6 | 2.1 |
| NCT00968708 | placebo | 65.0 | 62.3 | 2.7 |
| NCT01032629 | JNJ-28431754-100 mg | 63.3 | 61.8 | 1.5 |
| NCT01032629 | JNJ-28431754-300 mg | 63.9 | 62.5 | 1.4 |
| NCT01032629 | PLACEBO | 64.5 | 62.6 | 1.9 |
| NCT01131676 | BI 10773 10mg | 64.1 | 63.5 | 0.6 |
| NCT01131676 | BI 10773 25mg | 65.8 | 64.5 | 1.2 |
| NCT01131676 | Placebo | 67.5 | 65.1 | 2.4 |
| NCT01989754 | Canagliflozin | 66.6 | 63.6 | 3.0 |
| NCT01989754 | Placebo | 66.8 | 65.1 | 1.7 |
| NCT02465515 | ALBIGLUTIDE | 67.0 | 65.6 | 1.4 |
| NCT02465515 | PLACEBO | 66.9 | 64.8 | 2.1 |

#### Main treatment effects for HbA1c

The main treatment effects for each treatment versus placebo is shown in Figure S 2 for HbA1c for a standard network meta-analysis without covariates. The point estimates are the means and line-ranges indicate the 95% credible intervals. The plots are separated into mono-, dual- and triple-therapy. Red ink indicates fixed effects models and blue ink random effects models. The majority of treatments reduced HbA1c; the difference ranged from -0.5% to -1.5%. Figure S 3 is similar, but shows the main effect estimates for MACE.

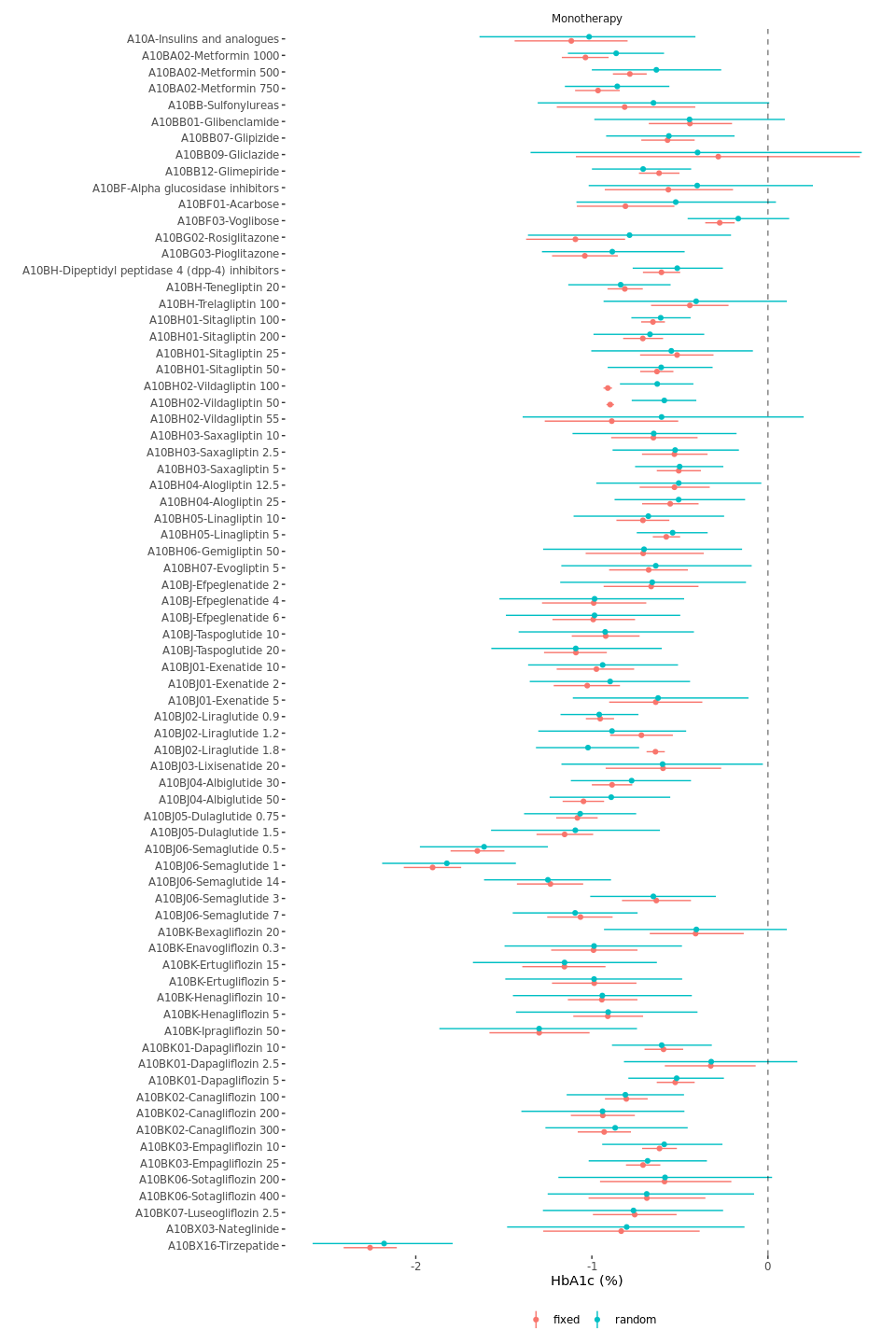

(a)

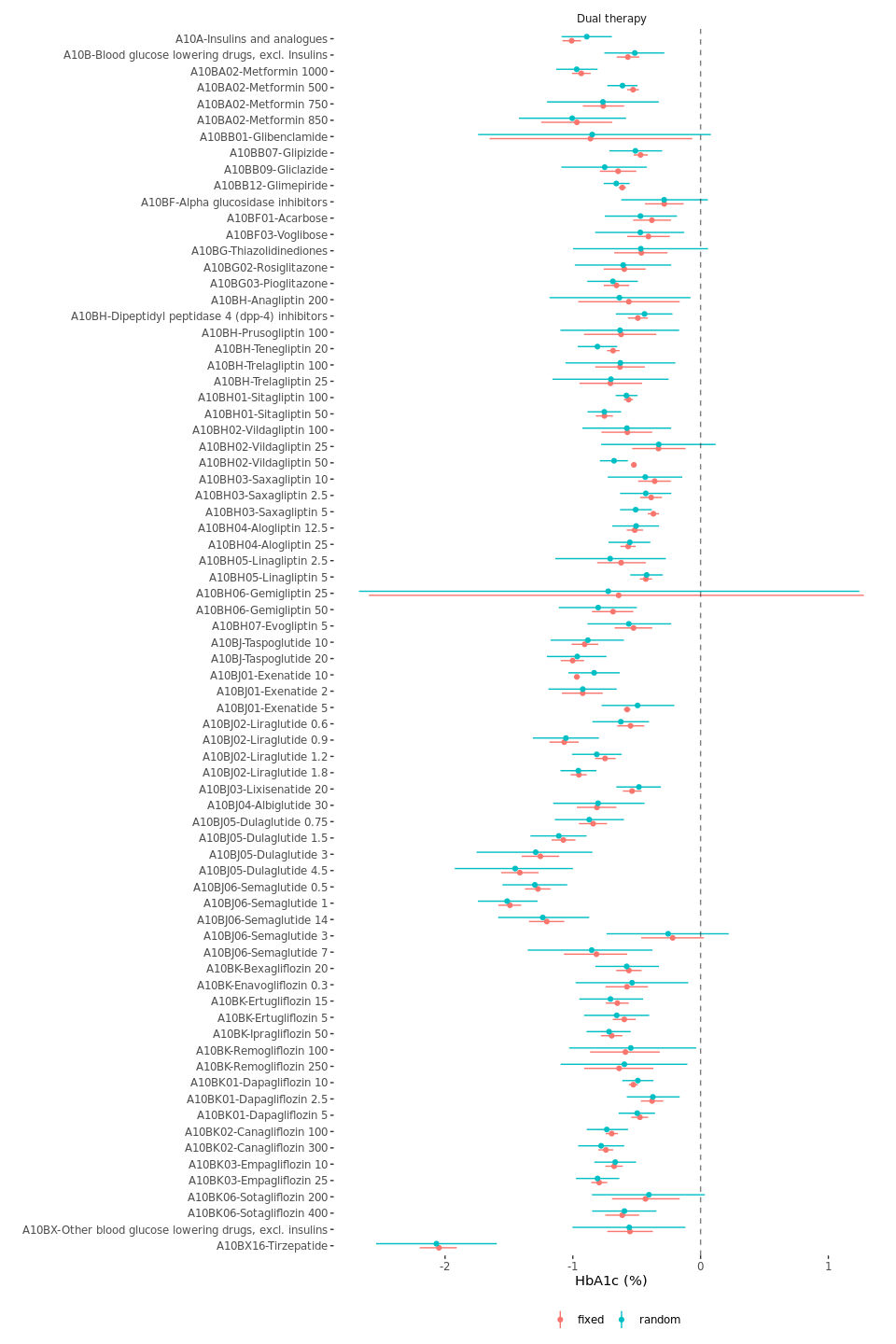

(b)

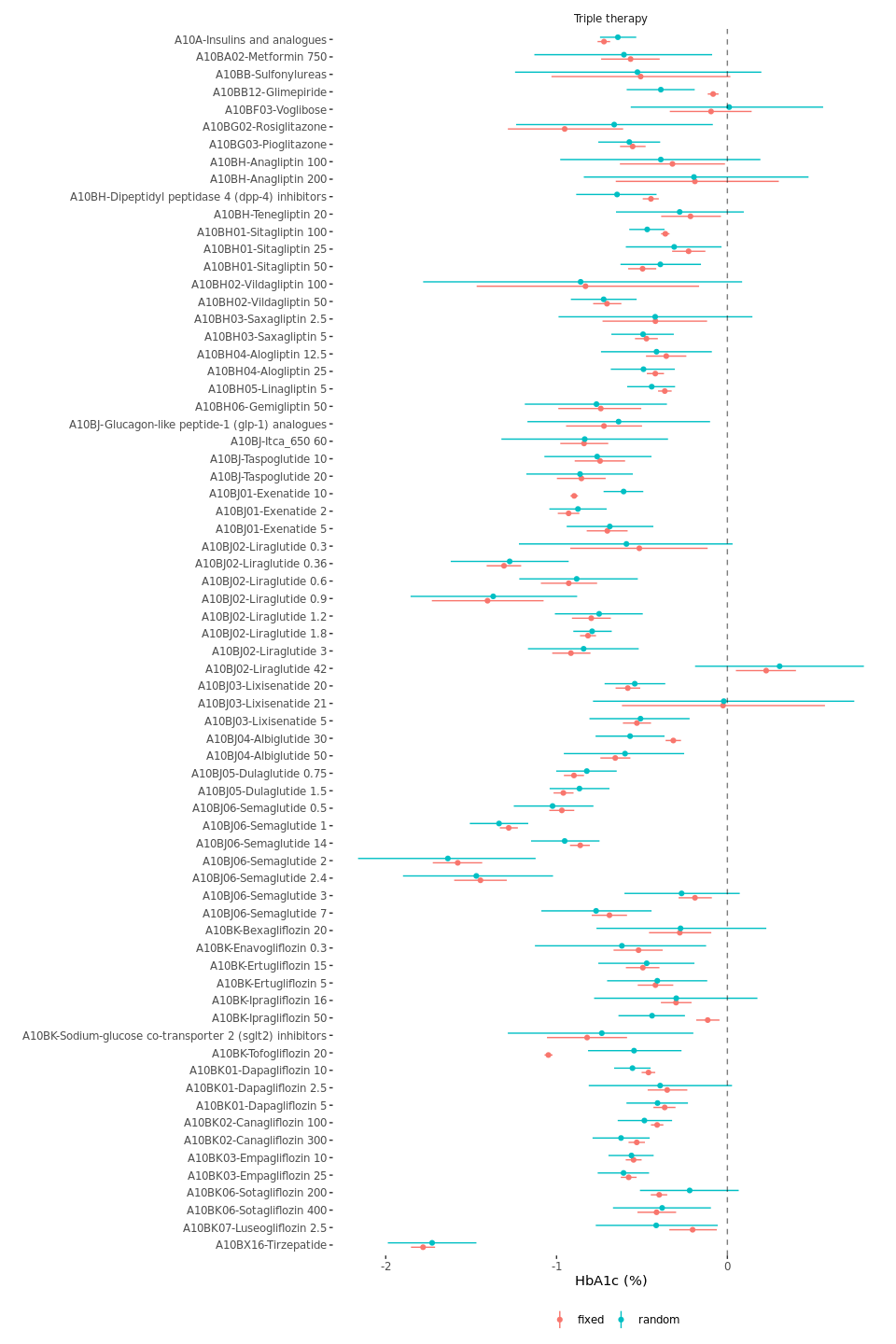

(c)

Figure S 2: Main treatment effects HbA1c

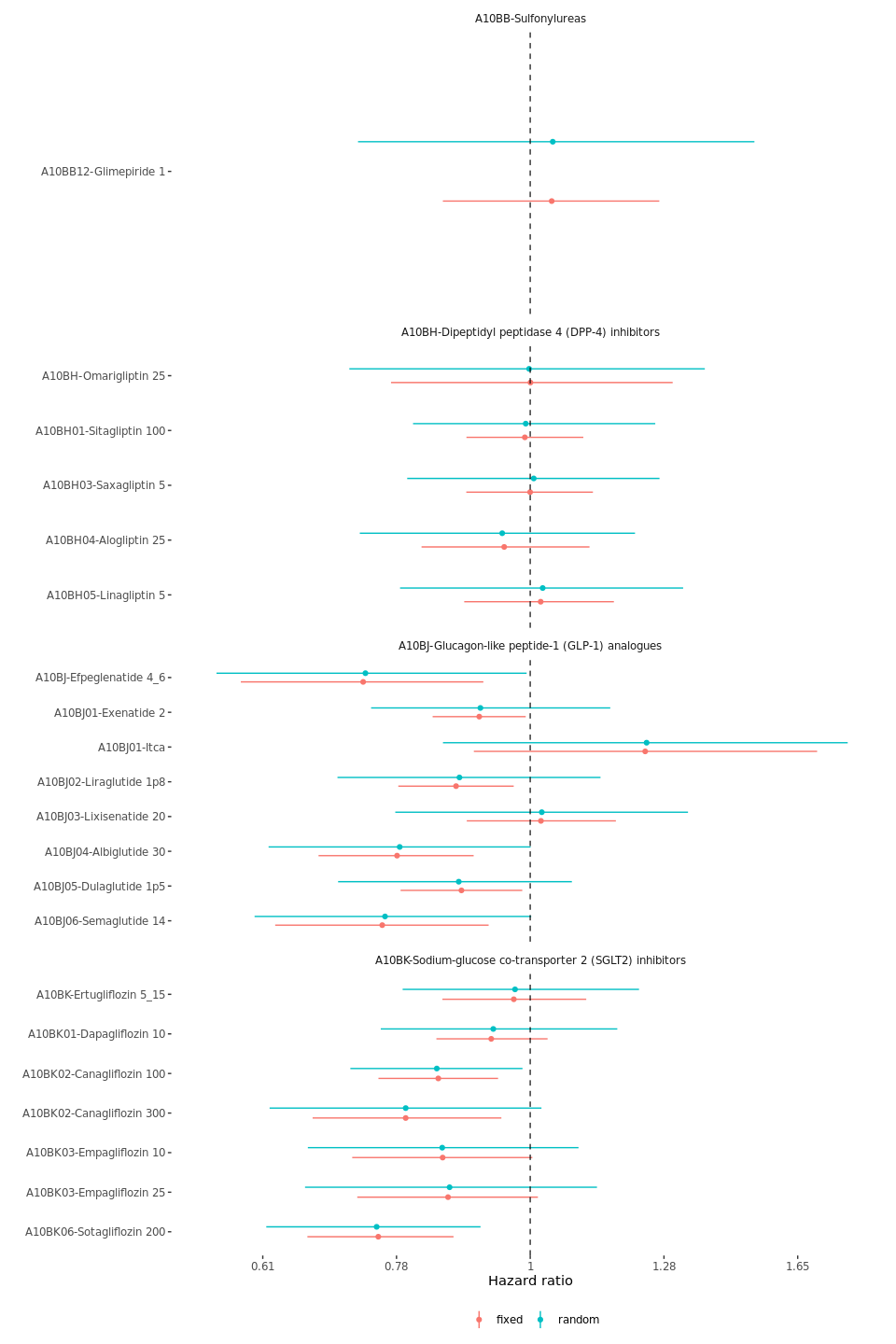

Figure S 3: Main treatment effects MACE

#### Sensitivity analyses with age- and sex-treatment interactions for MACE trials

Figure S 4 (a) and Figure S 4 (b) show the age-treatment and sex-treatment interactions for MACE for a range of sensitivity analyses respectively. The figures are panelled to show these interaction effect estimates for each drug class. As with Figure S 3, the points and line ranges represent the mean and 95% credible intervals of the posterior distribution. The plots show the findings for ‘all trials’ at the top of each panel. The use of subgroup data in the modelling is indicated using yellow ink, and the absence of subgroup data is indicated with black ink.

The bottom of the figures show the result of these analyses after dropping/downgrading selected IPD trials. Black ink indicates the results where the selected trial is completely dropped from the analysis, and no subgroup-level data is included in the analysis. Yellow ink indicates where the selected trial is completely dropped from the analysis, and subgroup-level data is included in the analysis where it is available for other trials. Blue ink indicates where the selected trial is not dropped but only downgraded from IPD-level to subgroup-level data and where subgroup-level data is included in the analysis where it is available for other trials.

We ran models dropping/downgrading (from IPD to subgroup data where this was available) for each of the 23 trials in turn. However, we only present results for each of the following three trials:- i) the single IPD trial for DPP-4s, ii) the single IPD trial for GLP-1s and iii) one of the four IPD trials for SGLT-2. We do so in order to simplify the presentation, as on dropping/downgrading each of the other trials there were no differences in the results when compared to the “all trials included” analyses (see model outputs in the project github repository - https://github.com/Type2DiabetesSystematicReview/nma_agesex_public).

All of these age-treatment and sex-treatment results were obtained using fixed effects models as it is not currently possible to fit random effects models with subgroup data in the multinma package. Nonetheless, similar results were found in fixed and random effects models for the models which did not include subgroup data (see main manuscript).

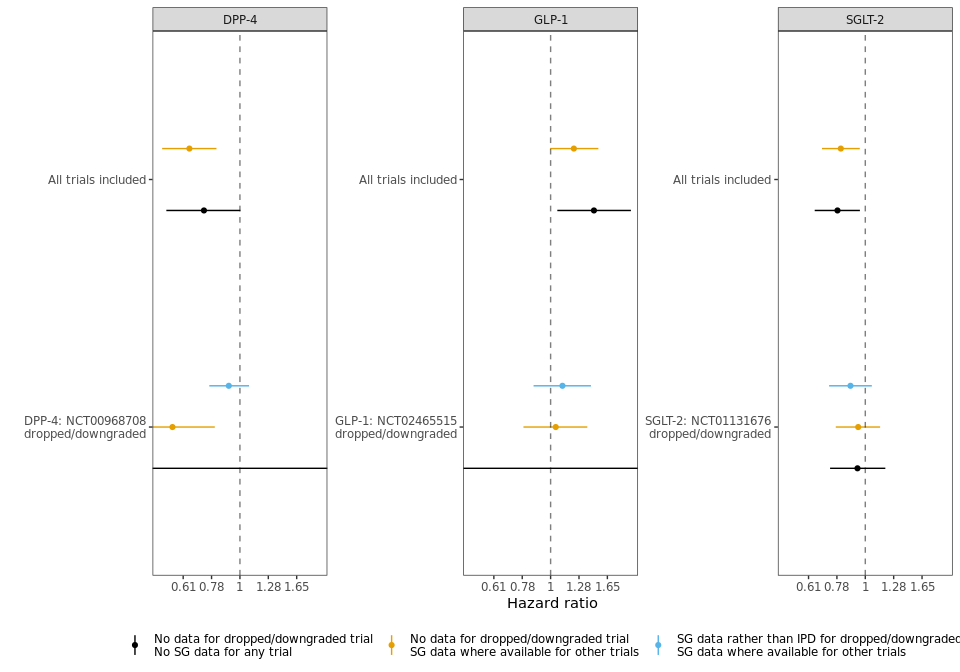

(a) Age interactions

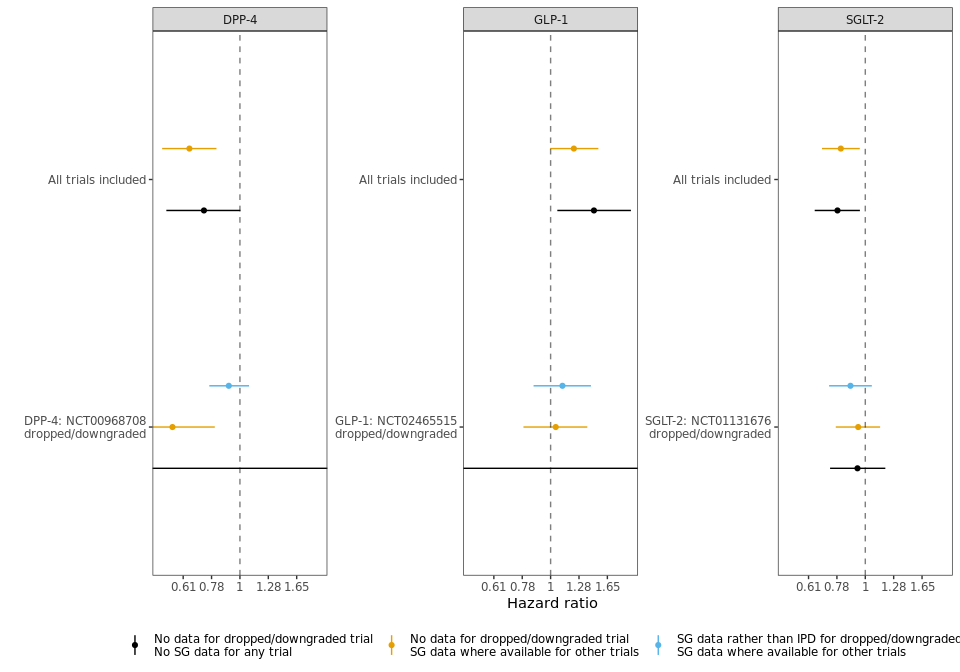

(b) Sex interactions

Figure S 4: Age and sex-treatment interactions for each class versus placebo
